# Clinical sequencing yield in epilepsy, autism spectrum disorder, and intellectual disability A systematic review and meta-analysis

**DOI:** 10.1101/2020.05.04.20089896

**Authors:** Arthur Stefanski, Yamile Calle-López, Costin Leu, Eduardo Pérez-Palma, Elia Pestana-Knight, Dennis Lal

## Abstract

**Importance:** Clinical genetic sequencing is frequently utilized to diagnose individuals with neurodevelopmental disorders (NDDs). Several reviews have been published regarding clinical genetic testing in various NDD subtypes. However, there is no systematic review and meta-analysis – in accordance with the PRISMA guidelines – which compares the genetic testing yield across neurodevelopmental disorder subtypes and sequencing technology.

**Objective:** To perform a meta-analysis and systematic review of the success rate (diagnostic yield) of clinical sequencing through NGS across NDDs.

**Data Sources:** Systematic review of the literature from PubMed until July 2019 for clinical sequencing studies that utilized NGS in individuals with epilepsy, autism spectrum disorder (ASD), or intellectual disability (ID).

**Study Selection:** Data were taken from clinical sequencing studies that screened more than five genes and performed variant classification in at least 20 individuals with epilepsy, ASD, or ID. 5.6% of identified studies met the selection criteria.

**Data Extraction and Synthesis:** Data were extracted, reviewed, and categorized according to PRISMA guidelines. Clinical evaluation and grouping were performed by two investigators following the ILAE guidelines. Pooled rates of the diagnostic yield and 95% confidence intervals were estimated with a random-effects model and adjusted for publication bias by the Duval and Tweedie procedure.

**Main Outcomes and Measures:** Diagnostic yield, defined as the proportion of individuals in a cohort who received a diagnosis based on a positive genetic test with variants identified as pathogenic or likely pathogenic.

**Results:** We identified 79 studies (epilepsy, n = 54; ASD, n = 13; ID, n = 17) across 29,301 individuals. Targeted gene panel sequencing was used in 53 cohorts and exome sequencing (ES) in 27 cohorts. The diagnostic yield was 16.7% for epilepsy, 20.2% for ASD, 24.8% for ID, and 16.6% overall. The diagnostic yield was significantly higher for exome sequencing compared to panels (33.9% vs. 16.2%, *P* = 1.38×10^−5^). We observed that the number of clinical sequencing studies increased annually, particularly studies from Asia (0-2 per year between 2012 and 2017, up to 10 in 2018). No studies from Africa, India, or Latin America were identified. We also found that recent studies are more likely to report variants of uncertain significance and few studies reported benign variants.

**Conclusions and Relevance:** This meta-analysis and systematic review provides a comprehensive overview of clinical sequencing studies of NDDs, which will help guide policymaking and steer decision-making in patient management.

**Key Points:** *Question:* What is the diagnostic yield of next-generation sequencing (NGS) in neurodevelopmental disorders and their subtypes?

*Findings:* In this systematic review and meta-analysis of 79 studies that include 29,301 individuals, the overall diagnostic yield was 16.6% (16.7% for epilepsy, 20.2% for ASD, and 24.8% for ID). Across all studies, downstream analyses showed a significant difference in yield between exome sequencing (33.9%) and targeted gene panels (16.2%).

*Meaning:* Around one in five NDD patients will receive a diagnosis using NGS, especially when investigating the whole exome.

## Introduction

Neurodevelopmental disorders (NDDs) – including epilepsy, autism spectrum disorder (ASD), and intellectual disability (ID) – represent genetically and clinically heterogeneous groups of disorders that affect about 3% of children worldwide.^1^ Advances in sequencing technologies have facilitated the identification of an exponentially growing number of NDD-associated genes.^2^ The identification of disease-associated genes can improve the understanding of disease pathogenesis and trajectories. It may also guide the identification of more genetically homogeneous subgroups within the spectrum of NDDs.^3^ A recent study showed that 33% of children with a molecularly confirmed genetic epilepsy would benefit from precision medicine.^4^ However, the proportion of individuals with NDDs who carry a genetic abnormality that can be identified using next-generation sequencing (NGS) has yet to be well established.

Sequencing studies that report the diagnostic yield (i.e., percentage of pathogenic variant carriers identified in a cohort) in NDDs are few but are becoming increasingly common. Estimates of diagnostic yield vary considerably across individual studies(8^5^–61%^6^). This likely reflects differences in measurement, reporting, and clinical characteristics such as etiology and disorder type/sub-type.

Although, many literature reviews have been published in the past, to the best of our knowledge, only two systematic meta-analyses of genetic testing in NDDs have been reported to date.

A literature review is a descriptive summary of the existing material relating to some topic or area of study. A systematic review is a review of the literature that is conducted in a methodical manner based on a pre-specified protocol and with the aim of synthesizing the retrieved information often by means of a meta-analysis. A systematic review sometimes produces results which, inconveniently, contradict common beliefs.^7^

A recent systematic meta-analysis of 30 NDD genetic testing studies showed that screening all genes with exome sequencing (ES) has a clinical diagnostic yield of 36% for individuals with NDD, 16% for a subset of individuals with ASD, and 39% for individuals with ID.^8^ Epilepsy was considered in only one systematic meta-analysis of genetic testing which focused on assessing the diagnostic yield of different technologies. The authors analyzed 23 epilepsy clinical genetic studies and found that ES had the highest diagnostic yield (45%; 6 studies), followed by targeted gene panel sequencing (panel) (23%; 9 studies), and chromosomal microarray (8%; 8 studies).^9^ Since NGS has only recently been established as a clinical diagnostic tool within the last decade, previous studies evaluating sequencing strategies have only included 30 studies or less. Furthermore, the diagnostic yield across subtypes of NDDs – including milder and more severe forms of epilepsy – or across sequencing technologies has yet to be consistently established.

Here, we present the most substantial and up-to-date systematic review and meta-analysis for clinical diagnostic sequencing in NDDs. In our study, we quantify the yield of diagnostic sequencing in different types of NDDs. We also explore sources of heterogeneity among studies and perform additional analyses considering the country of origin, type of sequencing test, and adherence to current variant interpretation guidelines.^10^

## Methods

### Search strategy

The systematic review was conducted following the Preferred Reporting Items for Systematic Review and Meta-Analysis protocol, considering all studies contained in PubMed until July 19, 2019.^11^ As keywords for the PubMed search, we used disease-specific terms (“epilepsy”, “epileptic encephalopathy”, “neurodevelopmental disorder”, “seizures”, “autism”, “ASD”, “autism spectrum disorder”, “intellectual disability”, “ID”, and “mental retardation”), each combined with sequencing technology terms (“exome”, “next generation sequencing”, “NGS”, “panel”, “targeted sequencing”, and “whole genome sequencing”) and other content related terms (“cohort”, “diagnostic yield”, “diagnostic test”, and “clinical practice”).

We performed an automated PubMed search using the R package RISmed.^12^ Moreover, 36 additional records were identified through other sources (e.g., listed as references in studies identified through PubMed screen). We only considered studies written in English. Duplicated studies, including response letters and studies without a title or abstract, were removed. The remaining studies were reviewed in two steps: 1) manual title and abstract screening to remove reviews, other non-original studies, and studies which did not perform clinical genetic testing using NGS technologies; 2) manual full-text review to select only sequencing studies which used NGS technologies, studies focused on germline variants, and studies that screened more than five genes in at least 20 individuals with epilepsy, ASD, or ID. We excluded studies that specifically ascertained individuals for congenital malformations of the brain or any other disorder where epilepsy, ASD, or ID were considered a secondary phenotype. The overall screening design is detailed in Figure 1.

**Figure 1.**
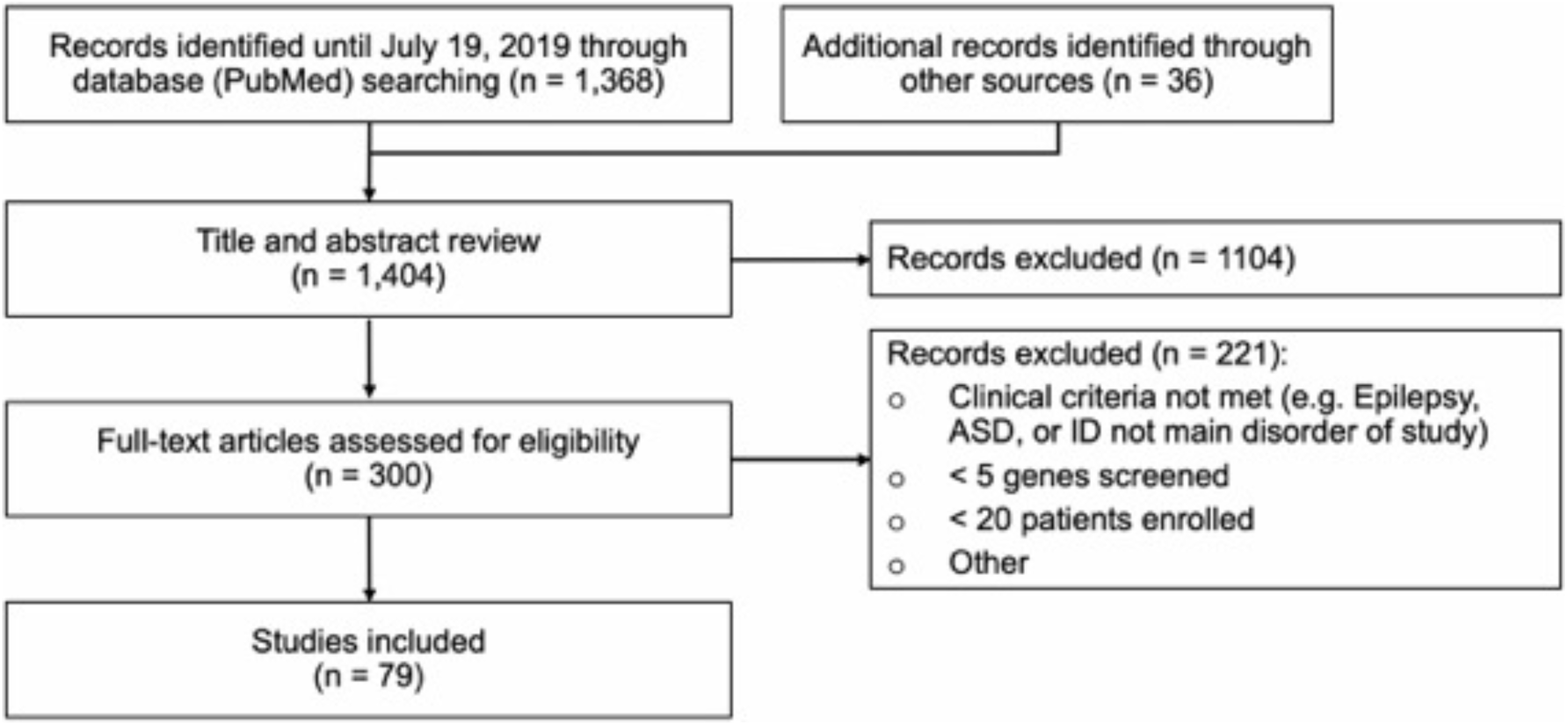
Process of data search, identification, and filtering

### Data synthesis and analysis

The 79 qualifying studies (eTable 1 in the Supplement) were divided into cohorts based on three criteria: 1) disorder, 2) disorder subtype, and 3) sequencing method. If a study investigated multiple disorders, disorder subtypes, or sequencing methods, we split these studies into disorder-specific, disorder subtype-specific, and method-specific cohort subsets (Figure 2). Disorder cohorts included: epilepsy, ASD, and ID. Disorder subtype cohorts included: generalized epilepsy (GE), focal epilepsy (FE), combined generalized and focal epilepsy (GE&FE), West syndrome (WS), and ASD with ID or developmental delay (DD). Method-related cohorts included: exome sequencing (ES) and targeted gene panel sequencing (panel). All cohort groups are detailed in Figure 2.

**Figure 2.**
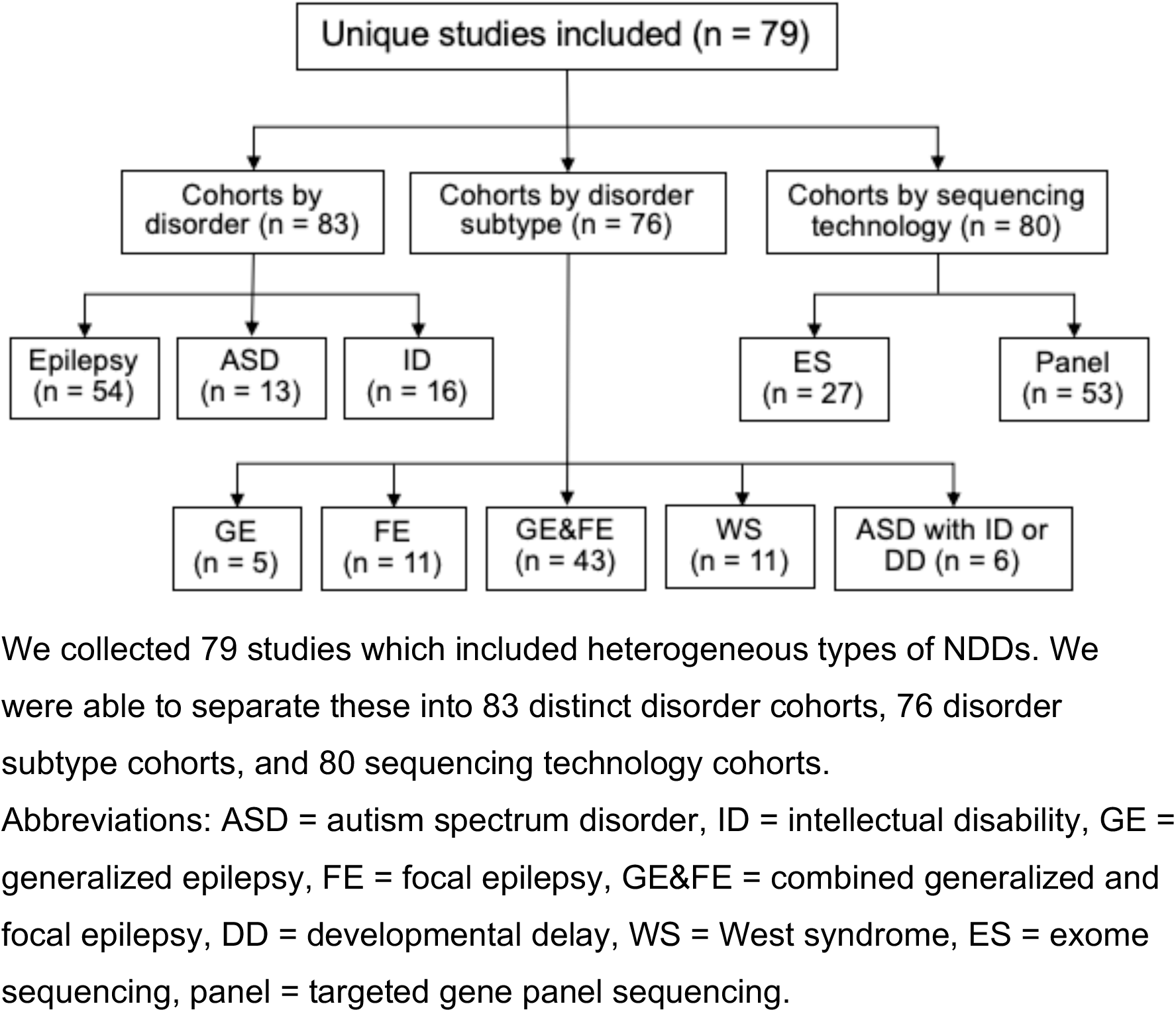
Separation of 79 unique studies included

**Figure 3.**
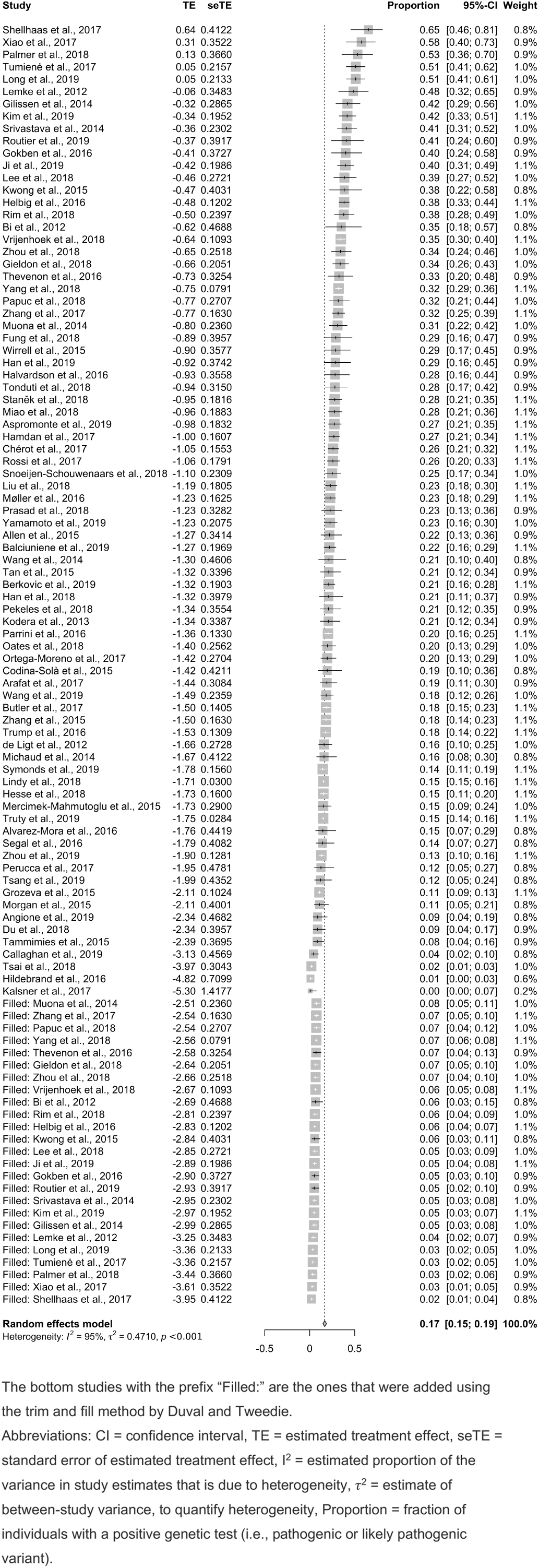
Forest plot of meta-analysis of the overall diagnostic yield from 79 studies

Small cohorts are more likely to be biased. Therefore, to enrich for representative studies, we did not consider studies that tested less than five genes or less than 20 individuals. Since they only reported coding variants, studies that employed whole-genome sequencing were included in the ES group. We only considered diagnostic yield from copy number variants (CNVs) if the variants were called from sequencing reads. Results from classical cytogenic testing were not considered.

Additionally, we assessed the number of studies that reported diagnostic sequencing across western (by continent: Australia, Europe, North America) and eastern countries (by continent: Asia) over time. We also investigated the number of disease-associated genes being reported by disorder per year. Lastly, we examined whether investigators applied the American College of Human Genetics & Genomics (ACMG) guidelines in NDD sequencing studies.

### Statistical analysis and statistical software

We used R version 3.6 for all the analyses.^13^ We performed systematic meta-analyses across all studies and cohorts (eFigures 1 to 23 in the Supplement) using a random-effects model (REM) with the R package meta.^14^ The REM was used considering an expected high degree of heterogeneity between studies. Plots were created using the meta and ggplot2 packages.^14,15^ The magnitude of between-study heterogeneity was quantified using the I^2^ statistic. A priori, we decided to report the pooled, weighted estimate generated by random-effects models, to account for a potentially high degree of between-study heterogeneity. We used funnel plots and the Egger method^16^ to evaluate potential publication bias. If bias was found, we performed a correction using the Duval and Tweedie trim and fill procedure.^17^ The Wilcoxon rank-sum test was used to determine significant differences between the diagnostic yield of panels and ES.

## Results

### Study selection

In our staged study selection process, we initially identified 1,368 unique studies through an automated PubMed search after inclusion and exclusion criteria were applied (Figure 1). Through other sources (e.g., reference lists), we identified 36 more studies. In total, we found 1,404 studies that met our criteria. We then eliminated 1,104 studies after abstract review, and another 221 after full-text review. A total of 79 studies, representing 29,301 individuals (mean: 371 ±1,439; median: 87, IQR: [48.5; 169]) were included in the systematic review. We conducted the analysis according to the Preferred Reporting Items for Systematic Reviews and Meta-Analyses (PRISMA) guidelines. The corresponding flowchart is shown in Figure 1.

### Diagnostic yield overall, by disorder and by disorder subtype

Using a random-effects meta-analysis of all 79 included studies revealed an overall diagnostic yield for neurodevelopmental disorder sequencing studies of 16.63% (95% CI: 15–19%) weighted by the number of cases in the study. Heterogeneity existed between estimates (I^2^ = 94.9%). In the disorder-specific analysis, the highest diagnostic yield was observed for ID (24.9%, 95% CI: 19–32%), followed by ASD (20.2%, 95% CI: 13–30%), and epilepsy, which had the lowest diagnostic yield (16.7%, 95% CI: 15–19%). Heterogeneity existed between estimates for ID (I^2^ = 92%), ASD (I^2^ = 89%), and epilepsy (I^2^ = 94%).

In the epilepsy subtype analysis, the diagnostic yield was 23.2% for GE (95% CI: 16–32%), 19.5% for FE (95% CI: 11–33%), and 16.4% for GE&FE (95% CI: 14–19%). Heterogeneity existed between estimates for GE (I^2^ = 84%), FE (I^2^ = 93%), and GE&FE (I^2^ = 95%). The highest diagnostic yields were observed for ASD with ID or DD (24.8%, 95% CI: 18–33%), and for West syndrome (WS; 24.4%, 95% CI: 16-35%) (eFigures 8 to 17 & 18 in the Supplement). Heterogeneity existed between estimates for ASD with ID or DD (I^2^ = 73%) and WS (I^2^ = 76%). All estimates were corrected for publication bias (see methods).

### Diagnostic yield by sequencing technology

Next, we stratified the study cohorts by sequencing technology (ES, n = 27; panels, n = 53). Random-effects meta-analysis showed a diagnostic yield of 33.9% for ES (95% CI: 29–40%) and 16.2% for panels (95% CI: 14–18%) (Table 1 & Figure 4). The difference was statistically significant (33.9% vs. 16.2%, 95% CI, *P* = 1.38×10^−5^). Heterogeneity existed between estimates for ES (I^2^ = 88%) and panels (I^2^ = 92%).

**Table 1.**
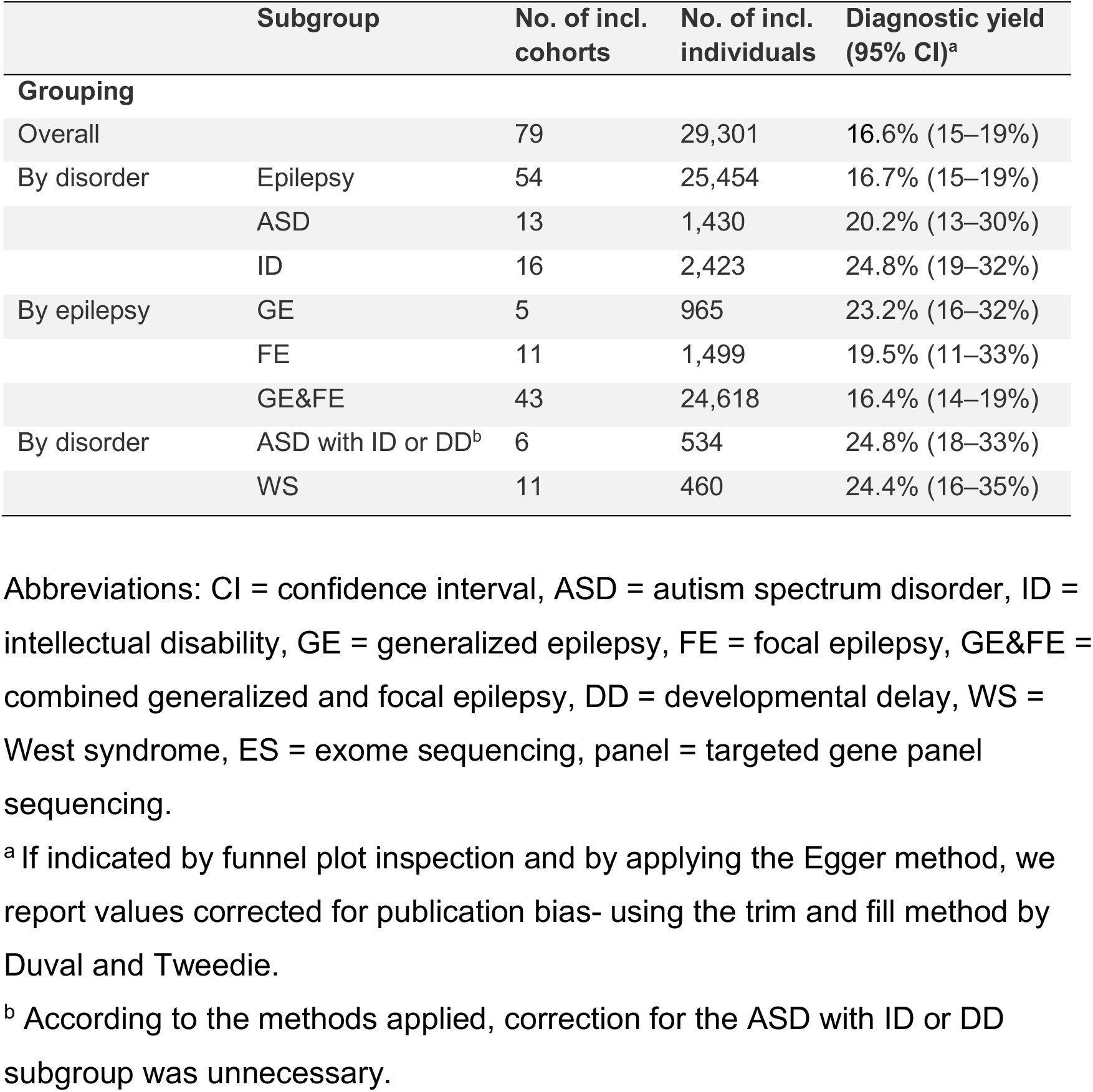
Diagnostic yield across different categories

**Figure 4:**
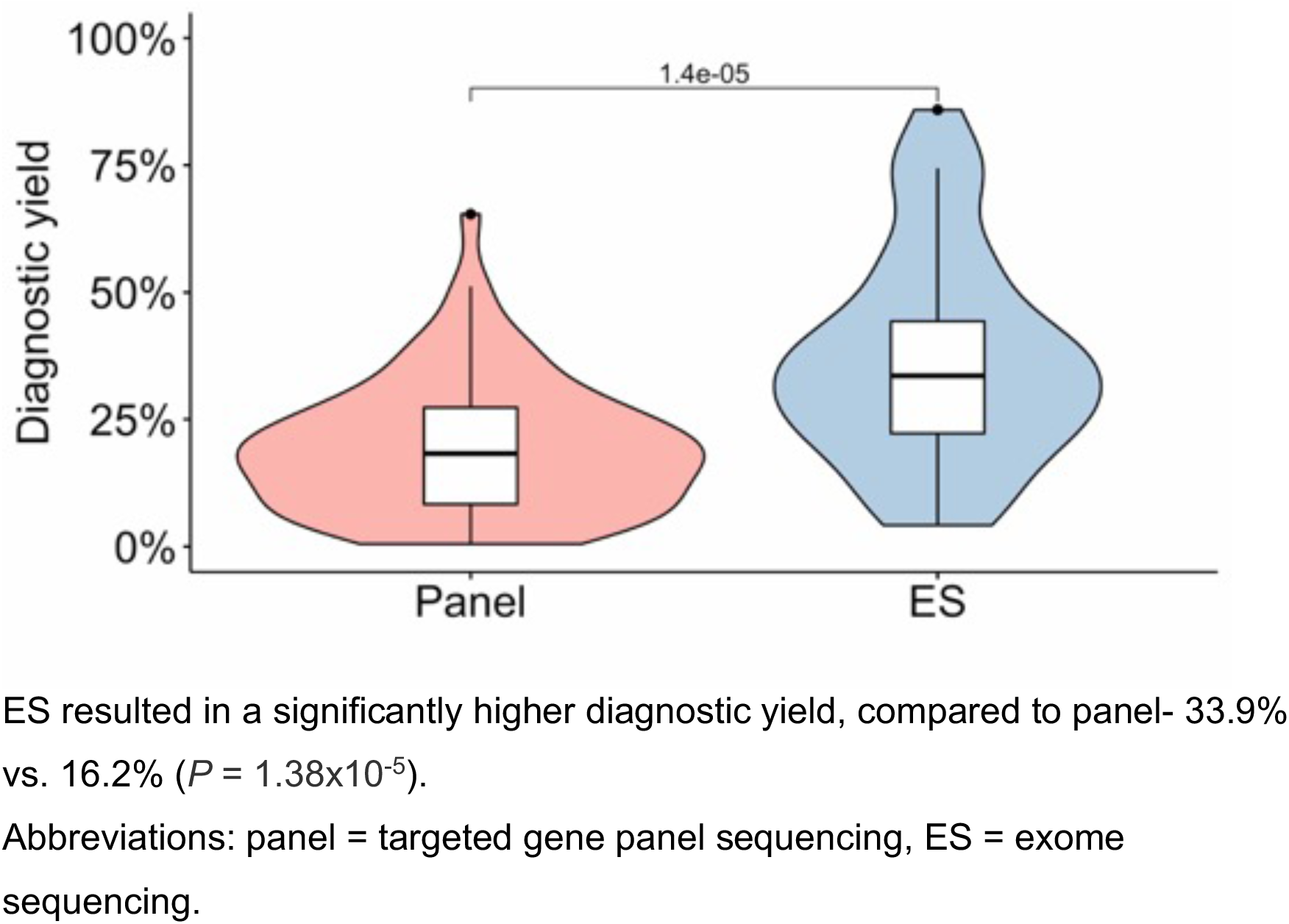
Diagnostic yield by sequencing technology

### Diagnostic sequencing utilization and variant interpretation over time

Diagnostic sequencing in neurodevelopmental disorders has only recently been introduced into clinical practice, and its adaptation across global regions has not been assessed. We assessed the number of studies reporting diagnostic sequencing across western (by continent: Australia, Europe, North America) and eastern countries (by continent: Asia) over time. Eastern countries, being exclusively from Asia, show rapid growth in publications in recent years (from 0-2 per year between 2012 and 2017, up to 10 in 2018). The study output from eastern countries increased from one study per year in 2012 and 2013 to one study per month in 2019, which is close to the output from western countries (~1.3 studies per month in 2019) (Figure 5). Between 2012 and 2017, 83% of studies originated in western countries, while eastern countries only produced 17%. In 2018 and 2019, western countries accounted for 55% of publications and eastern countries for 45%. Overall, 30% of studies were from Asia and 70% from western countries.

**Figure 5:**
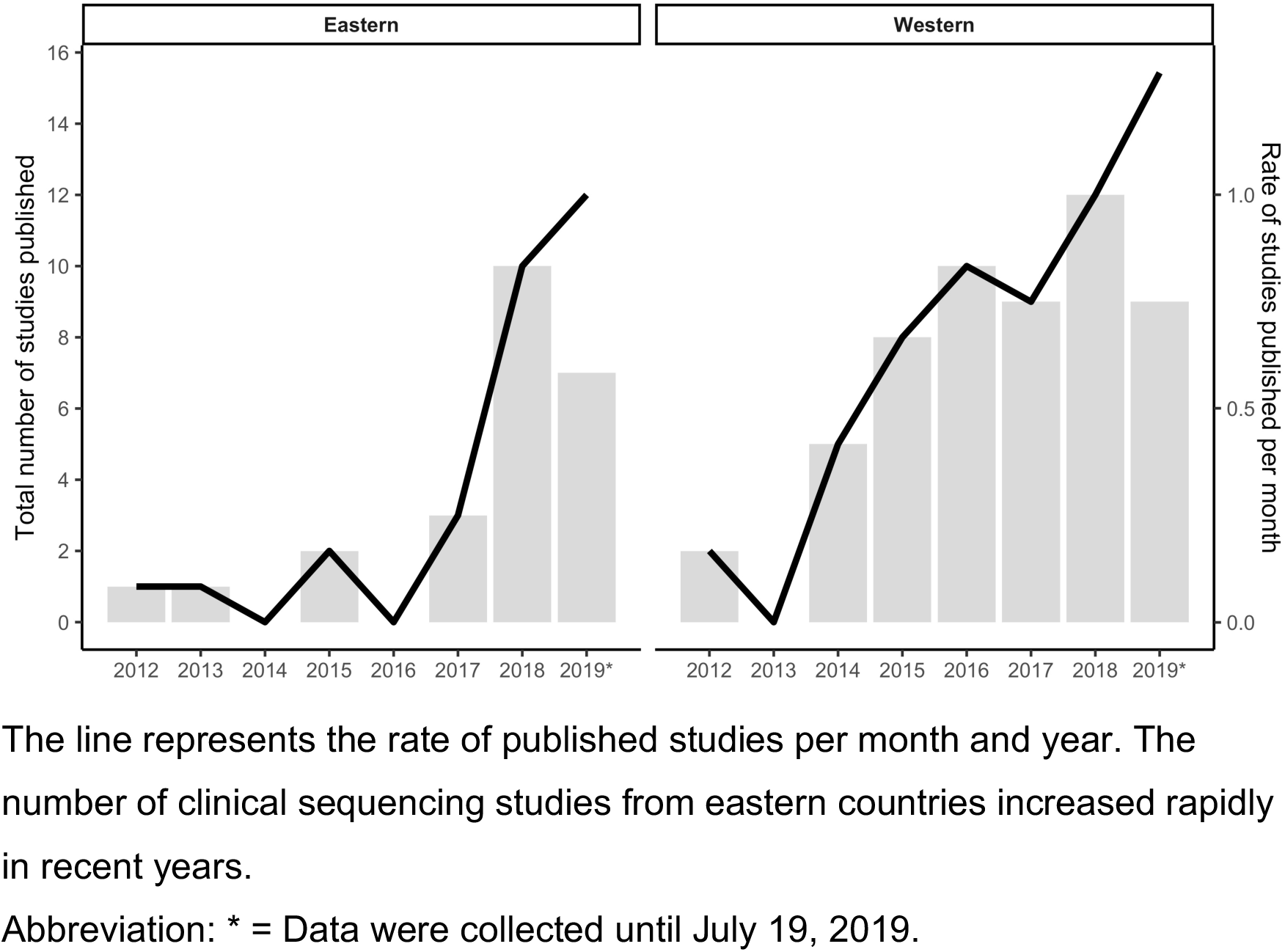
Total number of published studies per year separated for eastern and western countries

Furthermore, to demonstrate the continual discovery of new genes, we determined the number of genes being reported by disorder per year. We found an upward trend for the number of genes with pathogenic variants being reported (eFigure 24 in the Supplement).

However, not all genetic variants identified in a diagnostic test are pathogenic.^10,18^ Interpretation guidelines have been developed by the community, leading to the implementation of the (ACMG) guidelines in 2015. We examined whether investigators applied the guidelines in NDD sequencing studies. Most studies started to apply the ACMG guidelines in 2016, one year after the original publication (Figure 6 A). At the same time, we also observed an increase in studies that reported variants of uncertain significance (VUS), starting at around 20% of all reported variants in 2014 and stabilizing at around 50% in 2019 (Figure 6 B). Benign variants were only reported by a small fraction of studies. The percentage of studies reporting benign variants fluctuated from 0 up to 33% without any clear trend (Figure 6 C).

**Figure 6:**
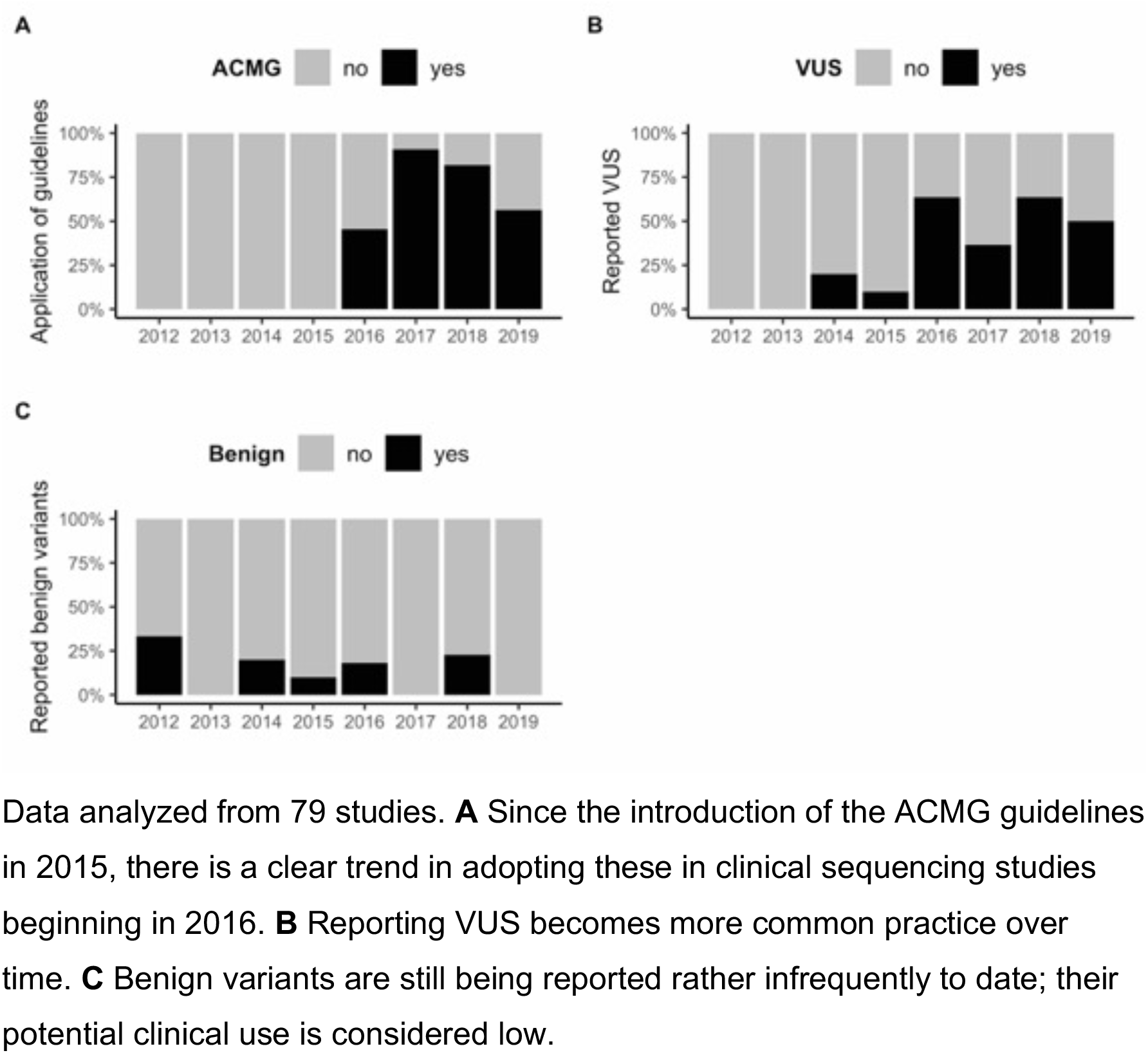
Level of variant interpretation and reporting as well as the proportion of the studies which reported VUS and benign variants

## Discussion

Here we present the largest systematic review and meta-analysis of clinical diagnostic sequencing in individuals with neurodevelopmental disorders. We identified 79 studies representing 29,301 individuals with epilepsy, ASD, or ID, and observed a cross-neurodevelopmental disorder diagnostic yield of 16.6%.

Our diagnostic yield of 20.2% for ASD, identified from 13 studies, corresponds to the diagnostic yield of 16% reported in a recent systematic meta-analysis of five studies.^8^ Our 16.7% combined diagnostic yield from panel and ES (n = 54 studies) for epilepsy is similar to 23% yield from panels (n = 9 studies) reported in a recent systematic meta-analysis.^9^ However, our combined diagnostic yield was lower than the 45% yield from ES (n = 6 studies) reported in the same study.^9^ Although the observed diagnostic yield of 24.8% for ID was the highest in our disorder analysis, the yield is lower compared to 39% previously reported.^8^ Additionally, recent articles report genetic testing diagnostic yields of up to 55-70% for individuals with ID.^19^

The study composition of this systematic review could explain the lower diagnostic yield for ID and epilepsy. Compared to the previous studies, the number of studies included in our systematic review was two to three times larger. Furthermore, we only included studies with at least 20 participants in order to increase statistical accuracy, which was not done by the previous systematic meta-analyses. This restriction could explain why our reported diagnostic yield is lower compared to other studies. Finally, we also included panel-based studies, whereas, the previous systematic meta-analysis on NDDs focused solely on ES data.^8^

In our sequencing method comparison, ES outperformed panel testing (33.9% vs. 16.2%, *P* = 1.38×10^−5^), consistent with two previous systematic meta-analyses.^89^ The yield for ES of 33.9% in our study is similar to a recent smaller systematic meta-analysis for NDDs without epilepsy, which reported a diagnostic yield of 31% for ES (n = 21 studies).^8^ Our systematic meta-analysis showed in between-study heterogeneity with I^2^ ranges from 88-95%, similar to previous systematic meta-analyses. In relation to the high I^2^ values, our results have to be interpreted with caution because diagnostic yields can vary widely across studies screening patients with apparently similar phenotypes. NDDs represent a clinically heterogeneous group of disorders, and differing patient ascertainment criteria could likely affect the diagnostic yield. Two studies labeling their cohort as “autism” cohort, could ascertain patients with different subtypes (e.g.; Asperger’ Syndrome vs. Pervasive developmental disorder) without indicating this information in the methods. The heterogeneity in the diagnostic yield of panel testing likely also reflects the different genes targeted by different panels.^2,20,21^

Several studies have successfully shown that guidelines are valuable in reanalyzing exome data.^22–26^ Before variant interpretation guidelines were developed five years ago, interpretation was not standardized. We show that, since their implementation, the field is progressively adapting the ACMG guidelines (Figure 3A).

Finally, we show a general trend of more NGS-based genetic testing worldwide with Asian countries showing the greatest increase in recent years. We did not find any study from Latin America, India, or Africa. Our data indicate that genetic testing is more common in western countries, and parts of Asia, compared to the rest of the world. We can only speculate that socioeconomic factors and a lack of genetics training could lead to this disparity.

This systematic meta-analysis should be interpreted in light of several limitations. First, diagnostic yield may be underestimated in some studies that pre-screened individuals and only performed NGS on patients for which a molecular diagnosis could not be established using standard genetic testing. Second, not all studies used the ACMG variant classification guidelines, which were first implemented in 2015.^10^ We also recognize that specific analysis approaches may differ in terms of variant filtering and technical platform (e.g., trio-based ES vs. proband-only ES). Additionally, the studies included in this comprehensive systematic review and meta-analysis represent a heterogeneous collection of sampling and data collection methodologies, with sparse descriptive information across all studies. Lastly, due to the absence of studies from Africa, India, or Latin America, the generalizability to individuals on a global level remains to be determined.

Nevertheless, this study represents the largest meta-analysis investigating diagnostic sequencing yield, including 2-3 times more studies than previous meta-analyses. In the absence of larger studies – excluding non-systematic reviews – this systematic review and meta-analysis can guide policymaking and help steer decision-making in patient management. Alongside policymakers and patients, healthcare providers can also benefit from this comprehensive overview. However, additional randomized controlled studies are still needed. Particularly, studies that examine the diagnostic determinants of optimal outcomes for children with rare genetic diseases.^27^

## Author contributions

AS and DL designed the study. AS, YCL and EPP analyzed the data. DL supervised the study. AS, CL, and DL wrote the manuscript. All authors interpreted the data and revised the manuscript.

## Data availability

All relevant data and methods are reported in the article and in the Supplement. **Study funding** No targeted funding reported.

## Disclosure

The authors report no disclosures.

## Glossary

ASD: autism spectrum disorder
CI: confidence interval
ES: exome sequencing
FE: focal epilepsy
GE: generalized epilepsy
ID: intellectual disability
NDDs: neurodevelopmental disorders
NGS: next-generation sequencing
panel: targeted gene panel sequencing
WS: west syndrome

## Data Availability

All relevant data and methods are reported in the article and in the Supplement.

